# COVID-19 IN IRAQ, THE RURAL INITIATION (AL-MUTHANNA PROVINCE AS AN EXAMPLE)

**DOI:** 10.1101/2021.03.16.21251969

**Authors:** Hazim Talib Thwiny, Safa Ibrahim Jaber, Hekmat Kadhum Ateya, Ali Mosa Rashid Al-Yasari, Nawar Jasim Alsalih, Moyed A. AL- Saadawe, Emad Salih Jasim, Mohenned A. Alsaadawi

## Abstract

A sustained pneumonia outbreak associated with a novel coronavirus named acute respiratory coronavirus 2 syndrome (SARS-CoV-2) was identified in Wuhan, Hubei Province, China in December 2019 which was later called COVID-19. The first confirmed case of COVID-19 was reported in Najaf/ Iraq on 24^th^ February. This paper provided some information on COVID-19 infection in the Province of Al-Muthanna / South Iraq, which was then statistically analyzed and concluded. Confirmed cases of COVID-19 infections were reported by the Iraqi Ministry of Health in the Province of Al-Muthanna. The first foci started and the first dead infected individual was from Hilal which refers mainly to the role or rural places in starting and transmission of COVID-19 in Iraq. Many of the infections resulted in non-traveling persons because they were contaminated by contact (96%). Therefore, contact is perceived to be the best-recognized form of transmission. It was also reported that infections in Soweir District of Samawah City were the highest (45%) compared to other areas of the region. They should also be observed, however, that the steps to enforce and monitor the curfew are directly related to the direction of the City Centre, because the more they drive away from the city centers, the less stringent the procedures. Infections were focused between the ages of 20 and 50 years old, as that is the expected result, because these ranges are at the core of active age groups including social and sports events.

**Graphical abstract:** 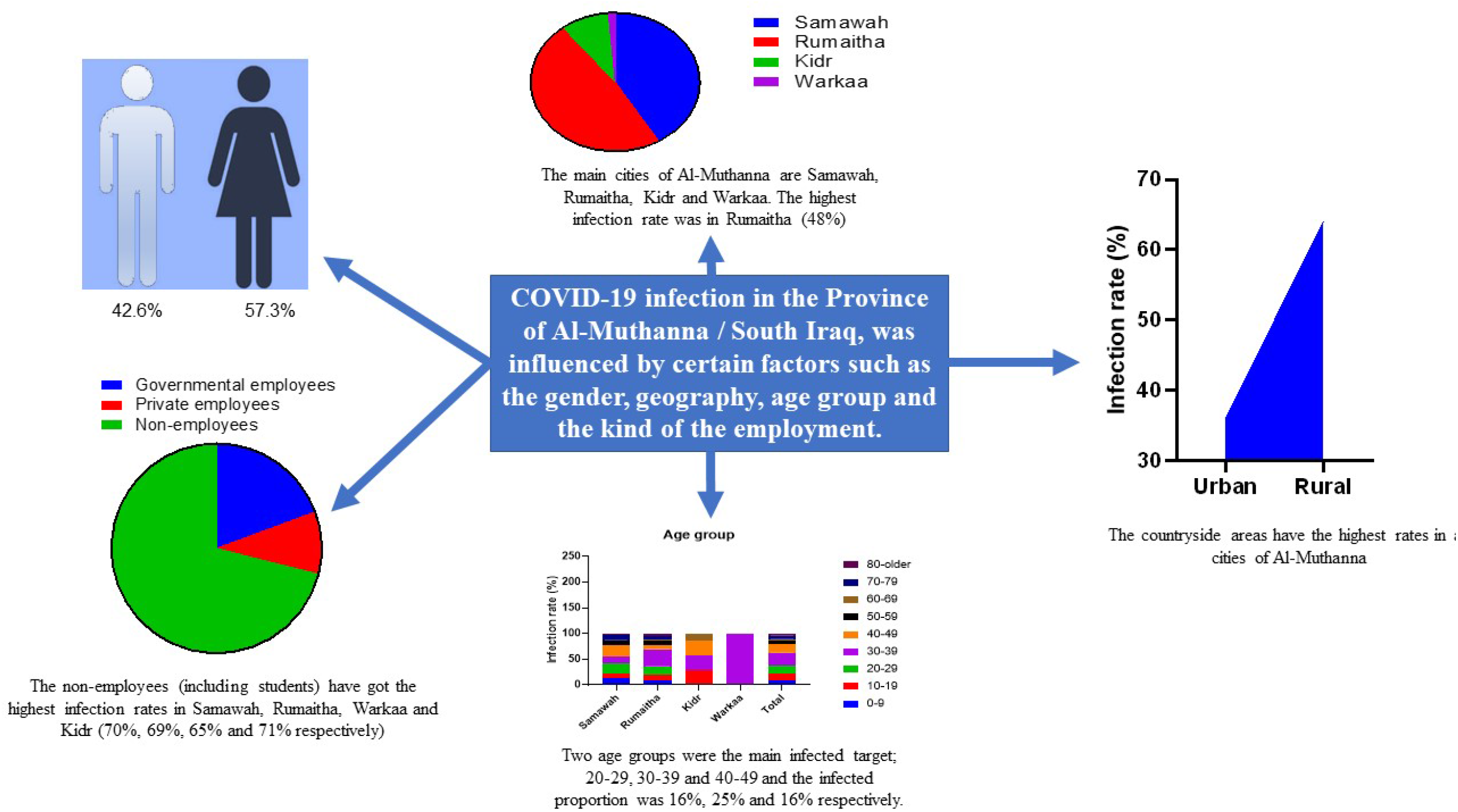

## INTRODUCTION

Coronavirus is one of the major pathogens targeted primarily at the human respiratory system. Previous coronavirus (CoVs) outbreaks include the Severe Acute Respiratory Syndrome (SARS)-CoV and the Middle East Respiratory Syndrome (MERS-CoV) previously identified as a major public health threat. An ongoing pneumonia outbreak associated with a novel coronavirus, named severe acute respiratory coronavirus 2 syndrome (SARS-CoV-2), was reported in December 2019 in Wuhan, Hubei Province, China [1–3]. Over the next few weeks, infections spread across China and elsewhere [4–6]. In order to allow the recognition and worldwide sharing of viral gene sequences, the Chinese public health, the clinical, and scientific community took immediate action [2,7] The outbreak was declared as a Public Health Emergency of International Concern (PHEIC) on 30 January 2020 by the World Health Organization (WHO) (World Health Organization, 2005). On 12 February 2020, WHO named Coronavirus Disease 2019 (COVID-19) [9] as the disease caused by the novel coronavirus. An international community of experts, with a number of specializations, has partnered with Chinese counterparts to find a way of the outbreak control [10].

The real-time reverse transcription – polymerase-chain-reaction (RT-PCR) assay for COVID-19 has been developed and used in clinics. Although RT-PCR remains the reference standard for definitive diagnosis of COVID-19 infection [11], the high false-negative rate [12] and the unavailability of RT-PCR assay at the early stage of the outbreak restricted early diagnosis of infected patients. Radiological examinations, especially thin-sliced chest CT, play an important role in the fight against this infectious disease. Chest CT can identify the early-stage of lung infection [13,14] [13,14] and prompt larger public health surveillance and response systems [15]. Chest CT results were currently recommended as key clinical diagnostic evidence in Hubei, China.

In Iraq, the first case of COVID-19 (also known as “coronavirus”) was reported in Najaf, 24^th^ February. Since then, 73 more cases have been confirmed, most of them in federal Iraq and around one-quarter of reported cases in the Kurdistan Zone of Iraq (KRI). Eight deaths were confirmed due to COVID-19. COVID-19 has been declared a major pandemic by the WHO. It indicates that the epidemic has officially spread around the world and represents the likelihood that the number of reported infections and fatalities will continue to increase. WHO has recognized that the dissemination of COVID-19 in Iran is now approaching the point of communal transmission, whereas the overwhelming majority of cases in Iraq are transmitted by people who traveled to Iran or who have been sent from Iran. WHO has reported that the infection in youth appears with comparatively milder symptoms and faster recovery periods (60% of the Iraqi population is under the age of 25), as the virus is not as dangerous for young persons as elderly individuals [16].

All “non-emergency” travel between the three KRI Provinces (Erbil, Sulaymaniyah and Duhok) has been forbidden by the Kurdistan Regional Government, which has declared that domestic flights between the Baghdad and Basra International Airports (14 March) and the Dutch International Airports (28 March) have been suspended. Jordan has forbidden traffic across the country from Iraq, however, flights run as normal [16]. The first foci started from rural regions of Rumaitha (Hilal district) according to The Ministry of Health which refers mainly to the role or rural places in starting and transmission of COVID-19 in Iraq. This paper will show some data of COVID-19 infection in Al-Muthanna Province/south of Iraq which then was statistically analyzed and concluded.

## DATA SOURCES

The confirmed COVID-19 cases were recorded by the Iraqi Ministry of Health for the COVID-19 infections in Al-Muthanna Province/ south of Iraq (Figure 1) and collected from the 8th of March till the 28th of April 2020. The cases were tested by the rapid IgG test and the results were confirmed in Central Health Laboratory (CHL) in Baghdad using real-time PCR. We quantified the number of new confirmed, recovered and dead COVID-19 cases per region of Al-Muthanna to find the peak of cases.

**Figure 1:**
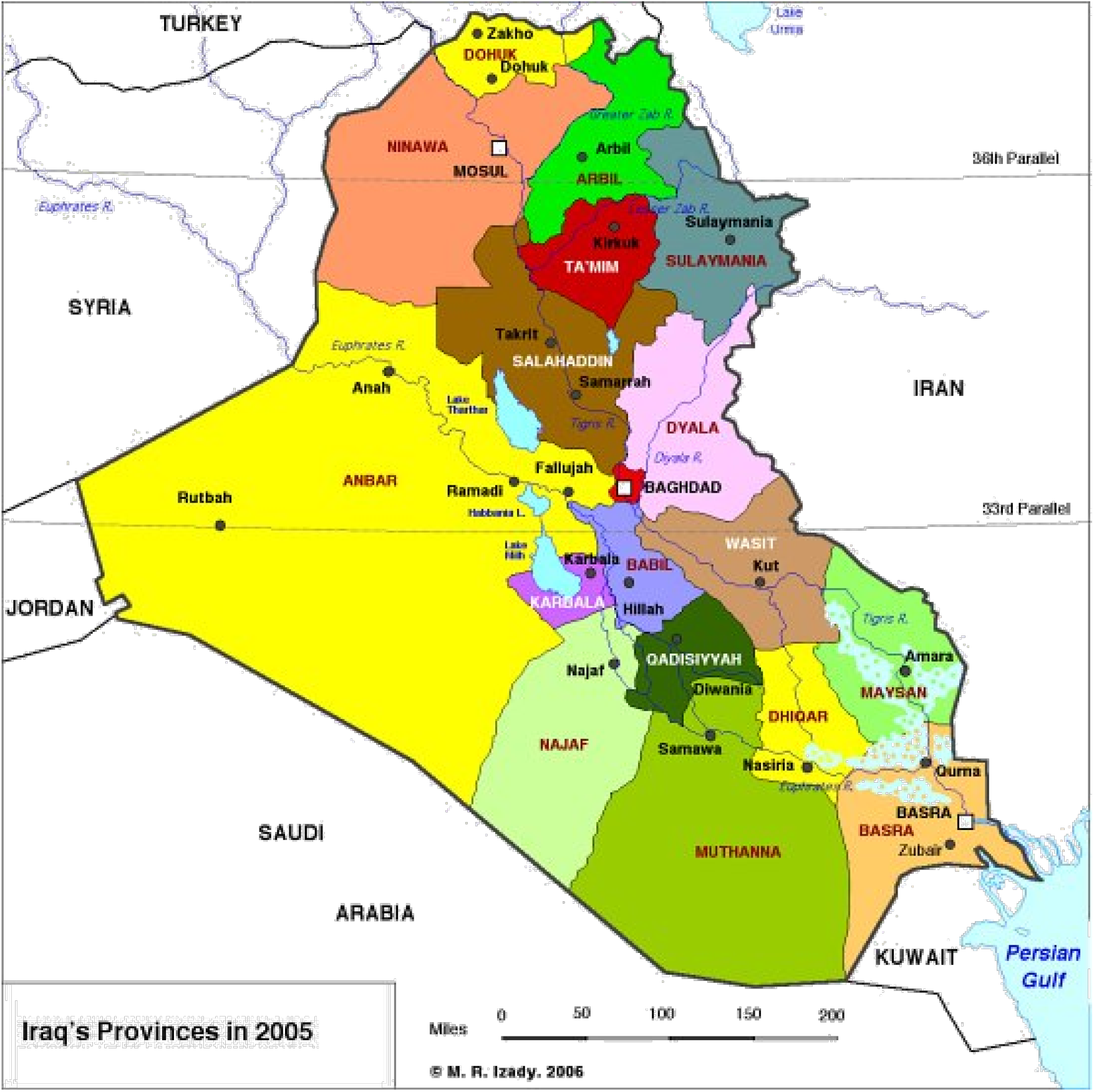
Iraq’s map includes Al-Muthanna Province (dark green) [17].

## RESULTS

The total infections with COVID-19 were 75 cases from the 8th of March till the 28th of April 2020 which can be separated into 72 (96%) patients have got the infection by direct contact while the other 3 patients (4%) were infected due to traveling to Iran in less than 2 weeks. The main cities of Al-Muthanna are Samawah, Rumaitha, Kidr and Warkaa. The highest infection rate was in Rumaitha (48%) (Figure 1 A). In this study, rural areas were more infected than urban in all tested cases (Figure 1 B). The infection rate in Samawah Districts showed some differences (Figure 3 A). The infection was found mainly in Soweir (45%) which is the largest countryside region of the city. Other places of Samawah center were also showed infections with varied rates. While in Rumaitha, most infections were in Hilal District (69%) (Figure 3 B). Still, the countryside areas have the highest rates in both the bigger cities of Al-Muthanna. The statistical analysis showed that there is a significant difference between males and females infected with COVID-19 (Figure 4 A). The nature of the job plays a role in the transmission ofCOVID-19. The non-employees (including students) have got the highest infection rates in Samawah, Rumaitha, Warkaa and Kidr (70%, 69%, 65% and 71% respectively) (Figure 4 B). Two age groups were the main infected target; 20-29, 30-39 and 40-49 and the infected proportion was 16%, 25% and 16% respectively. These age groups showed significant differences (Figure 5).

**Figure 2:**
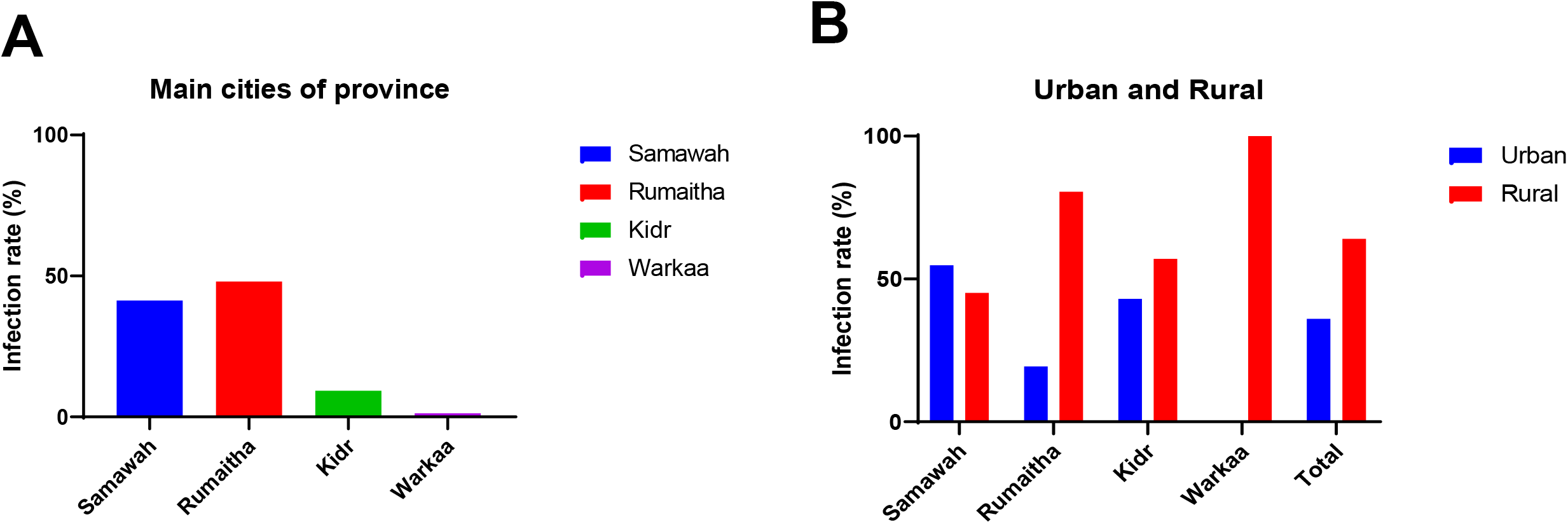
Infection rates of the main regions of Al-Muthanna Province. Data of infection in the main cities of Al-Muthanna were recorded (A). The rural places in each of these cities were more infected than urban (B). Statistically significant differences were done for each column using t test, P <0.0001 between the rural and urban areas of the main cities of Al-Muthanna Province.

**Figure 3:**
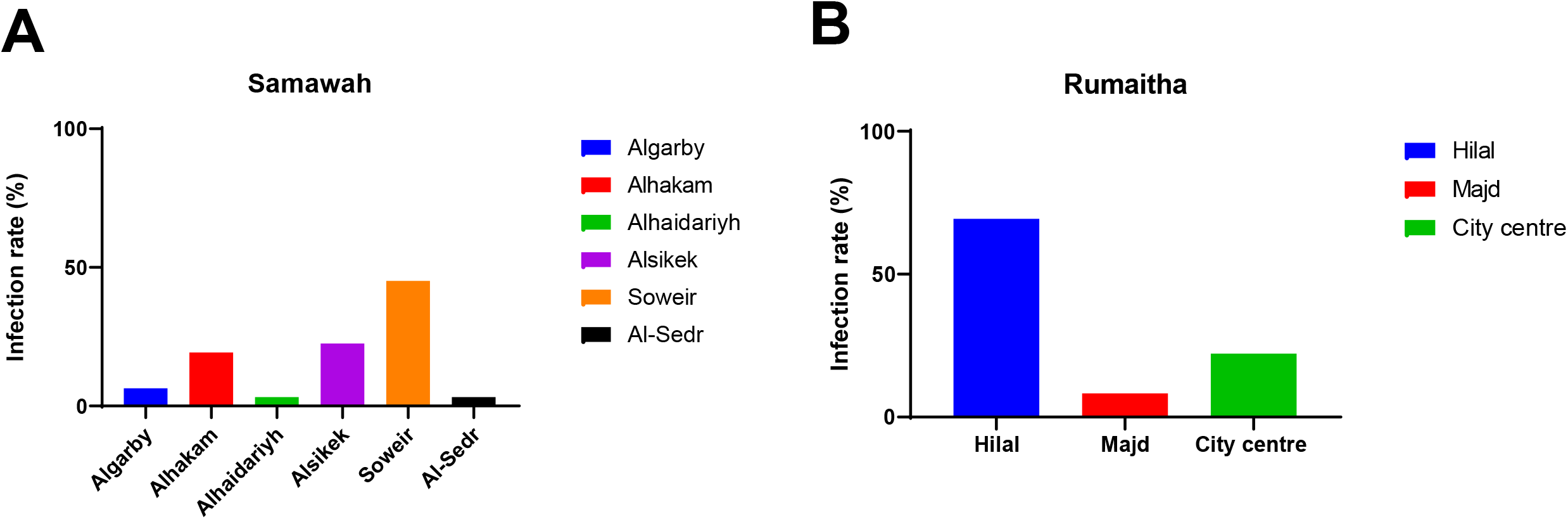
Infection rates of the main regions of Samawah and Rumaitha. The countryside places of Samawah showed high percentage of infection (Soweir) (A) and the same thing found in Rumaitha as the highest rates were in Hilal District (B).

**Figure 4:**
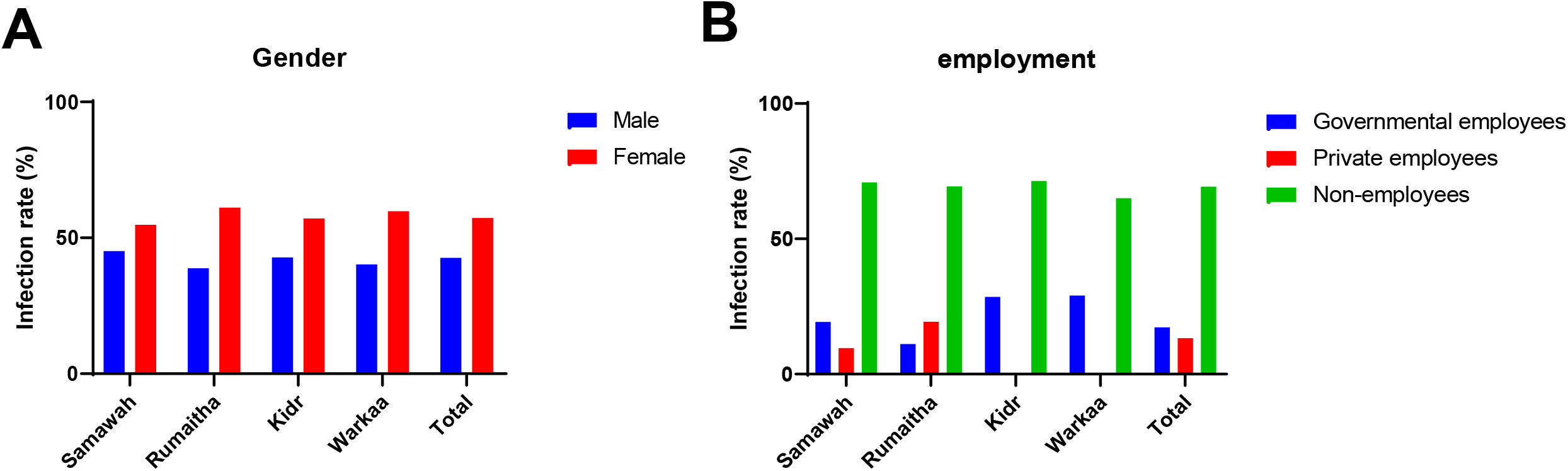
the infection rates according to the gender and work. The rates of infection were classified into male and female infections (A). Infections were also classified according to kind of jobs (B). Statistically significant differences were done for each column using One-way ANOVA and Tukey’s multiple comparisons test, P <0.0001 between males and females and between governmental, private and non-employed infected individuals of the main cities of Al-Muthanna Province.

**Figure 5:**
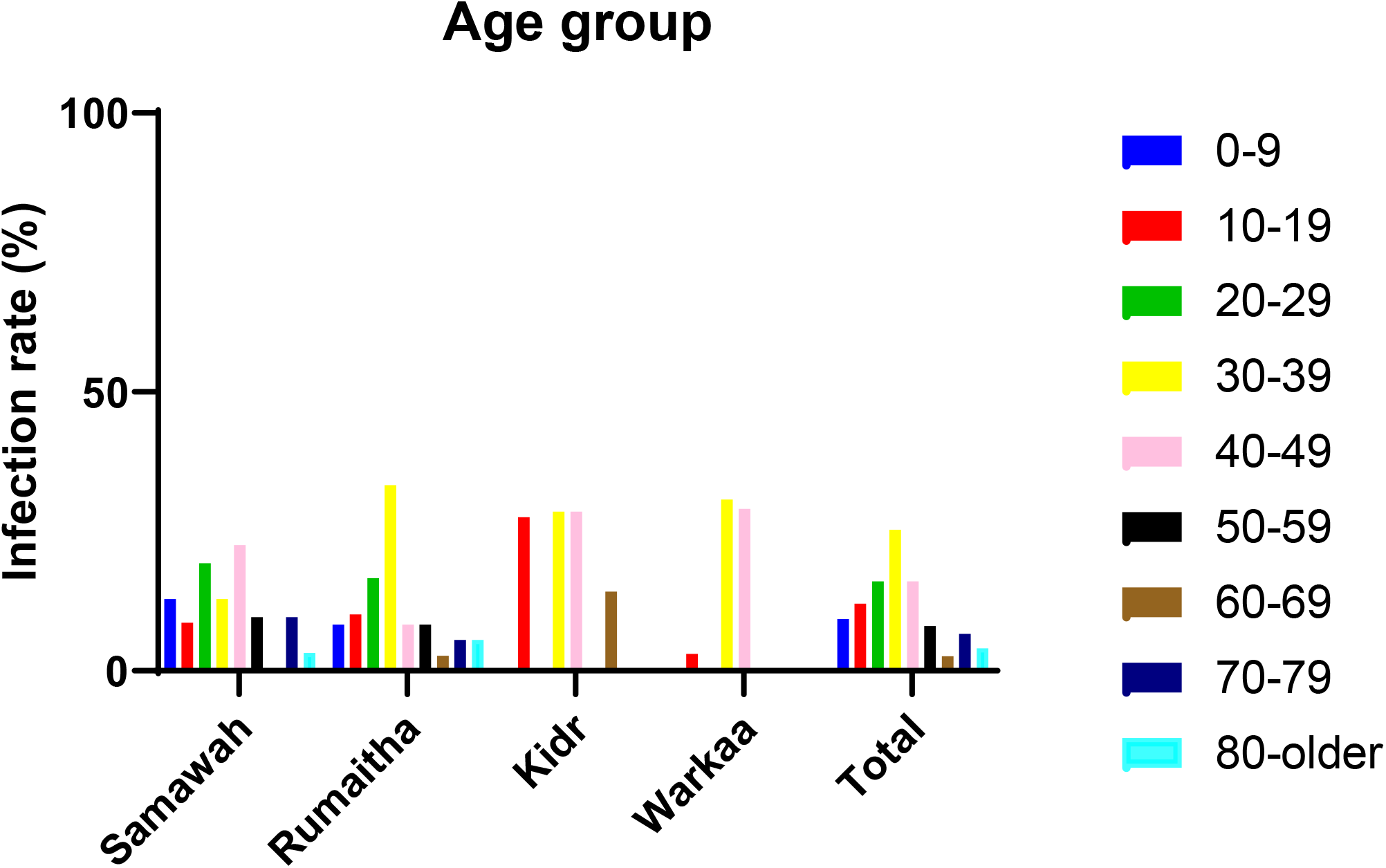
the infection rates according to age groups. Different age groups were targeted by COVID-19 virus. Two age groups were got the highest infection rates which are 30-39 and 40-49. Statistically significant differences were done for each column using One-way ANOVA and Tukey’s multiple comparisons test, P <0.0001 between the different age groups of the main cities of Al-Muthanna Province.

## DISCUSSION

Most of the infections were for non-traveling individuals, as they get infected by contact (96%). Therefore, the contact is considered to be the most early known transmitted way than other methods [3,18]. There is no evidence that the arrivals from endemic countries increase the infection rates as the most Iraqis came in from abroad outside of Iraq were free of the disease. The highest proportion of infection was recorded in Rumaitha city and its Districts (48%), especially in Hilal, where the infection rate reached 69% of the total cases that appeared in Rumaitha. The main possible cause associated with the highest infection rates in Hilal could be the nature of the region which is rural which is considered as a persisting challenges of initiation of infection [19]. The implementation of curfews in this kind of region is difficult as the curfew could be one of the main methods to decrease the daily infection rates of COVID-19 [20]..

Likewise, it was noted that the infections in Soweir District of Samawah city (the capital of Al-Muthanna Province) were the highest (45%) compared to the other areas of the city. Also, the measures implementation the curfew could be the reason, as they were completely applied in city centers and main streets only unlike the Districts and rural areas and this is clear from the total infection rates in the countryside. Moreover, the lack of health awareness about the risks and methods of COVid-19 transmission that must be followed to prevent the disease can also be an important cause of the increased infection rates as the awareness of COVID-19 symptoms are main predictors of preventive actions [21,22]. Hence, we have also noticed that the measures to implement and monitor the curfew are directly proportional to the approach of the city center, as the further we move away from the city centers, the less strict the procedures.

The infections focused in the ages of 20-50 years, and this is an expected result, as these ages represent the core of working age groups and have social and sports activities that need gathering frequently. Within these age groups, it was found that the non-employed represent the highest infected people. Health awareness and following the instructions of the Ministry of Health, especially with regard to how to avoid the disease, have an important role and no less than the importance of curfews. We, therefore, recommend the following:

1. More roles should be given to Health Crisis Committee and their team.
2. Encouraging people to commit to social distance and preventing kissing, cuddling, speaking for close distances, and touching the face, mouth, nose, and crowding in shops for shopping, especially during the partial curfew.
3. The curfew, as implement on cars, must be imposed on pedestrians since direct contact is the main transmission factor for viruses. The movement of pedestrians without masks is strictly prohibited.
4. Forcing shop owners to place signs that define social distances to leave a safe distance between the shoppers.
5. Allow researchers to easily access all information about COVID-19 and the methods used for diagnosis and treatment for the scientific and statistical evaluation.
6. Establishing an advanced virology laboratory containing modern equipment, especially PCR, and with experts and collaborated with specialists from the University of Al-Muthanna. The goal of establishing this kind of lab is to quickly diagnose and genetic sequencing of viral infections.

## Conclusion

One of the main pathogens in the human respiratory system is a coronavirus. In Iraq, the first case of COVID-19 was recorded in Najaf on 24 February. WHO has reported that the infection appears in young people with comparatively mild symptoms and faster recovery times. Most of the infections occurred in non-traveling individuals as they were infected by contact (96%). Therefore, communication is perceived to be the best recognized form of transmitting. The highest proportion of infection was registered in Rumaitha and its Districts, especially in Hilal. It has also been reported that infections are higher in Soweir than in other parts of the Samawah. The curfew policy enforcement should be applied since it is not strictly implemented in Districts and rural regions, they were entirely implemented in the city centers and main streets, and that is evident from the higher rates of infection in the rural areas. Furthermore, a significant factor that can involve in transmission which is the lack of awareness information on the hazards and strategies of Covid-19 transmission. Infections have been concentrated from 20 and 50 years old of age. This is an expected outcome, since these groups are the main working classes and have social and sports activities that need gathering frequently. It has been found within these age groups, the most infected are the non-employees. Health education is an essential role in the transmission and not less than the value of curfews.

## Data Availability

The data that support the findings of this study are openly available in zenodo https://zenodo.org/badge/DOI/10.5281/zenodo.4548253.svg

## Acknowledgments

We all thank the Iraqi Health Ministry which facilitates and provides us accurate details about the initiation of COVID-19 in Al-Muthanna province. Also, all the appreciation for Veterinary Medicine Collage/ Department of Microbiology which established the research group to help in the pandemic control.

---

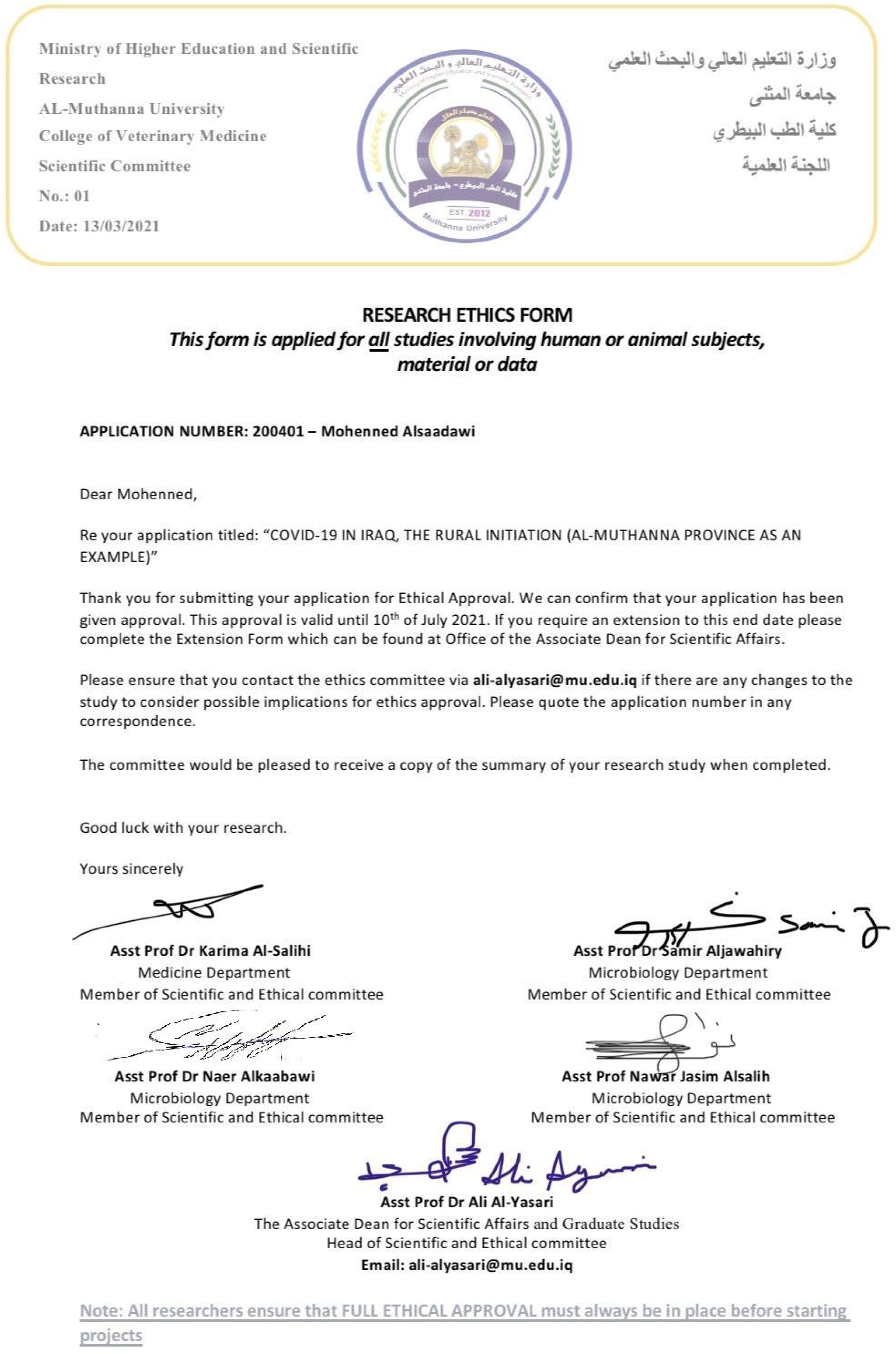

